# Longitudinal associations between depressive symptoms and brain structure across late childhood and adolescence: A panel network analysis study

**DOI:** 10.64898/2026.03.24.26349162

**Authors:** Eira R. Aksnes, Dani Beck, Niamh MacSweeney, Marieke G.N. Bos, Lia Ferschmann, Linn B. Norbom, Valerie Karl, Lars T. Westlye, Christian K. Tamnes

## Abstract

**Background:** Major depressive disorder (MDD) is the leading cause of non-fatal disability in youth and disproportionately affects adolescent females. Structural MRI studies of adolescent depression have yielded inconsistent findings, potentially reflecting symptom heterogeneity and rapid developmental changes in brain morphology.

**Methods:** We examined associations between specific depressive symptoms and structural brain MRI measures in 9,761 youth (53.1% male, age range = 8.32–18.0) from the Adolescent Brain Cognitive Development (ABCD) Study across four waves. A panel graphical vector autoregression (GVAR) model separated within-person (contemporaneous and temporal) from between-person effects. Brain measures included cortical thickness in four regions, hippocampal volume, and global surface area. Depressive symptoms included parent-reported depressed mood, anhedonia, lethargy, worthlessness, and sleep problems. Sex differences in network structure were also examined.

**Results:** Strong within-domain associations were observed among brain measures and symptoms. Cross-domain associations emerged at both within-person and between-person levels. Within persons, higher depressed mood predicted subsequent decreases in global surface area, whereas higher worthlessness predicted subsequent increases; depressed mood was also linked to mOFC thickness over time and to lower global surface area concurrently. At the between-person level, individuals with higher average anhedonia showed lower average global surface area. Sex differences were confined to the within-person networks.

**Conclusions:** Brain-symptom associations were subtle, symptom-specific and present across network levels with differential edge stability. Global surface area emerged as the primary structural correlate. Longitudinal within-person approaches capture the coupling of within-person deviations from expected brain and symptom trajectories, complementing between-person approaches in understanding brain-depression co-fluctuation during adolescence.

## INTRODUCTION

Depressive disorders have become the leading cause of disease burden in adolescence, with incident rates rising sharply across this developmental period, particularly among adolescent females (1–3). Adolescent-onset MDD is associated with high rates of comorbid mental disorders and somatic conditions (4–6), and an early onset predicts a more recurrent and severe illness course into adulthood (7–9). Protracted neurodevelopmental change, including widespread reductions in grey matter volume, cortical thickness, and surface area, alongside heterogeneous subcortical volume changes characterizes the adolescent period (10–14). Given that depression rates rise sharply in this same period, there is considerable interest in whether deviations from typical brain development confer depression risk (15).

Cross-sectional studies report subtle structural brain differences in adolescent depression, though findings vary by metrics and sample types. Global cortical surface area reductions have been observed in clinical MDD samples (16,17), while both cortical volume and surface area reductions have been associated with depressive symptoms in community-based samples, including in ABCD (18).

Longitudinal evidence from community samples has linked childhood depressive symptoms to accelerated prefrontal thinning and volumetric loss across adolescence (19), while accelerated prefrontal thinning has in turn been associated with greater concurrent depressive symptoms in late adolescence (20). However, no such associations emerged in high-risk or clinical MDD samples, whether examining depression predicting later cortical thickness (21–23), or co-occurring trajectories of symptoms and brain structure (22). In contrast, illness onset in high familial risk samples was associated with cortical thickening in inferior frontal regions, rather than thinning (24). Subcortical findings are mixed, with some studies linking hippocampal and amygdala volume change to depression outcomes (24–26). Whether well-established sex-differences in depression prevalence (1,3,27) are parallelled by sex differences in brain–depression associations remains unclear and under-examined, with some studies identifying female sex as a predictor of depression onset via neurobiological pathways and others finding no such effect (23,28–30).

Previous inconsistent findings may partly reflect how depression has been operationalized across studies. While several studies have adopted dimensional approaches by examining depressive symptoms continuously rather than categorically (18,20,31), these have largely aggregated symptoms into total scores. This aggregation may obscure neural correlates specific to individual symptoms rather than overall severity (32) and masks the substantial heterogeneity in symptom profiles and their dynamics that can differ substantially even at equivalent levels of severity (33,34). Studies also differ in whether they examine concurrent or prospective brain-depression associations, in sample characteristics (community vs. clinical vs. high risk), and in the specific brain regions and metrics examined, all of which may contribute to inconsistency across findings.

The network approach to psychopathology addresses these limitations directly by reconceptualizing depressive symptoms not as indicators of a latent disorder, but as a complex system of interacting components that mutually influence one another over time (35,36). Critically, this approach moves beyond total symptom scores by modelling associations at the level of individual symptoms and their neural substrates across development (37). Adolescence is a particularly compelling context for this approach, given that concurrent change in brain structure and depressive symptoms suggests bidirectional interplay between neurodevelopment and symptoms (8,38,39). Symptom-level investigation is further warranted in community samples, where depressive symptoms frequently fall below diagnostic thresholds (40).

Only a few cross-sectional studies have taken this approach. In an adult sample with a history of depression and healthy controls, one study reported symptom-specific associations with brain regions most consistently implicated in adult MDD (41). This included associations between smaller hippocampal volume and sadness and loss of interest and between thinner cortex in the insula, cingulate, and fusiform gyrus with distinct affective and cognitive symptoms. Extending this approach to older adolescents, another study found that thinner cortex in the cingulate and thickening in the insula were associated with worthlessness, thinner mOFC was associated with anhedonia, and larger hippocampal volume with sleep problems (42). Critically, associations were absent when depression was operationalized as an aggregate severity score, demonstrating that symptom-level operationalization can reveal associations that aggregate approaches miss. Differing association patterns across these two studies suggest developmental variation. Moreover, as both studies were cross-sectional and did not examine sex differences, questions remain regarding the temporal dynamics of brain-symptom associations across adolescence and whether these are sex-specific.

The present study addresses these gaps by employing a network approach to a large, longitudinal four-wave sample of youth (N = 9,761, 53.1% male, age range = 8.32–18.0) to examine how depressive symptoms relate to brain structure across late childhood and adolescence. We applied panel network analysis to capture dynamic, symptom-level associations between depressive symptoms and brain structure over time, while separating within- and between-person processes. Following previous work (41,42), we focused on regional cortical thickness and hippocampal volume to extend this symptom-level framework to an earlier developmental window and a longitudinal design. Global cortical surface area was additionally included given its consistent association with depressive problems across both clinical MDD samples (16,17) and community-based adolescent samples (18).

Based on prior network studies (41,43) we hypothesized that thinner cortex in the cingulate would be associated with increased worthlessness and sadness and that thinner mOFC would be associated with increased anhedonia. We also hypothesized that hippocampal volume would be associated negatively with sadness and positively with sleep problems. Given preliminary evidence that brain-depression associations may not be uniform across sexes (23,28,31) we examined sex-differences in brain-symptom associations as an exploratory aim. Finally, we examined whether brain-symptom associations differed when depressive symptoms were modelled individually compared to when aggregated into a sum score.

## METHODS AND MATERIALS

### Sample and Ethical Approval

We used longitudinal parent-reported symptoms and MRI data from the Adolescent Brain Cognitive Development (ABCD) Study (44). Participants were excluded according to the ABCD Study criteria, outlined in the 7.0 data release notes and briefly summarized in SI Section “ABCD Inclusion/Exclusion criteria”. The Institutional Review Board at the University of California San Diego approved all aspects of the ABCD Study (45). Caregivers provided written consent, while the child provided written assent. The present study was conducted in line with the Declaration of Helsinki and approved by the Norwegian Regional Committee for Medical and Health Research Ethics (REK 2019/943).

### Demographic Information and Data Quality Assurance

The final sample included 9,761 unique participants (9,556 at TP1, 6,558 at TP2, 5,153 at TP3, and 6,167 at TP4), contributing 27,434 observations across one or more of four timepoints (baseline, year 2, year 4, and year 6) with concurrent T1-weighted MRI and symptom data. To address family-level clustering inherent to the ABCD Study design, one participant per family was randomly selected (fixed random seed) and retained, removing 2,031 siblings and twins. See Supplemental Information (SI; Section MRI Acquisition and Processing) for further details. The sample was predominantly White (63%); full race/ethnicity distributions are reported in Table S1. The number of participants contributing observations declined across waves, reflecting MRI related attrition (Figure S1). Sample demographics are reported in Table S2. A descriptive summary of depressive symptoms and brain features can be found in Table S3 and S4, while mean trajectories are shown in Figure S4. Age distributions across the study period are illustrated in Figure 1.

### MRI Acquisition and Processing

Neuroimaging data were acquired across 21 different sites using 31 scanners (3T Siemens Prisma, GE 750, or Philips systems) and processed centrally by the ABCD Study Team (44). Acquisition, processing protocols and quality control procedures are described in detail elsewhere (44,46). Following recommended ABCD data quality assurance, we additionally filtered cases using the provided QC variable (mr_y_qc incl smri t1_indicator). In the present study, we extracted T1-weighted imaging (T1) tabulated regional measures of cortical thickness in the cingulate, mOFC, insula, and fusiform gyrus, as well as hippocampal volume and global surface area (SI Section “MRI Acquisition and Processing”). Global cortical surface area was included as an additional brain node as it represents the most consistently implicated structural metric in adolescent depression across large-scale samples, with reductions observed in both clinical and community-based cohorts including ABCD (15,16,18). Imaging data were harmonized across scanners using the *long.Combat* R package (47,48), specifying sex, timepoint and CBCL items used in the main analysis as covariates, to preserve variance associated with these variables. In a following step, left and right hemisphere values were averaged, and hippocampal volume was residualized by intracranial volume and sex following exact procedures of previous symptom-brain network studies (41,43). Detailed descriptions of these procedures can be found in SI Section: Long.Combat Harmonization.

### Depression Symptoms

To capture core depressive symptoms aligned with the DSM-5 criteria for MDD, we selected five parent-reported items from the DSM-oriented Depressive Problems scale of the Child Behavioral Checklist (CBCL) (49,50): depressed mood (“Unhappy, sad or depressed”), anhedonia (“There is very little he/she enjoys”), worthlessness (“Feels worthless or inferior”), lethargy (“Underactive, slow moving or lacks energy“) and sleep problems (“Has trouble sleeping”). Following the principle of minimally complete node set (51), items were selected to maximize coverage of distinct non-overlapping DSM-5 depressive symptom dimensions, prioritizing symptoms particularly relevant to adolescent depression (52) and largely consistent with prior symptom brain network studies (41,53). Depressed mood and anhedonia represent the two core DSM-5 MDD criteria (54), while worthlessness and lethargy capture cognitive-evaluative and psychomotor dimensions of adolescent depression, and sleep problems cover the somatic dimension of adolescent depression. Items were rated on a three-point scale (0 = Not True; 1 = Somewhat/Sometimes True; 2 = Very True or Often True), reflecting behavior within the past six months.

### Statistical Analysis

All analyses were conducted using R version 4.3.2 (R Core Team, 2024). To examine associations between specific depressive symptoms and selected brain measures over time, we estimated a panel graphical vector-autoregression (GVAR) network model using the *psychonetrics* package (56) applied to four-wave panel data. The panel GVAR separates processes that unfold within individuals across time and stable between-person differences (57). However, the panel GVAR allows a larger number of continuous variables to be modelled simultaneously and estimates multiple network structures within a unified framework (51,58).

The panel GVAR model decomposes associations into three complementary network structures (53,59,60). In all networks, depressive symptoms and brain measures are represented as nodes and edges reflect pairwise partial correlations, representing the unique association between two variables after accounting for all others in the model. The *temporal network* models directed lagged effects, capturing whether deviations from an individual’s mean in one variable predict deviations in another variable or the same variable (autoregressive effects) at the subsequent wave. The *contemporaneous network* captures within-person co-deviation associations within the same measurement window, after accounting for temporal and between-person effects. The *between-person* network captures associations between individualś averaged levels across time, reflecting stable trait-like differences. Models were estimated using full information maximum likelihood (FIML), retaining participants with incomplete data.

The model assumes stationarity, such that network structures are assumed invariant across the modeled developmental period. To facilitate stationarity, all variables were examined for linear and quadratic time trends and detrended accordingly prior to model estimation (58). This standard procedure in panel modelling removes systematic developmental trends, allowing the temporal network to capture within-person deviations around individual means, the contemporaneous network to reflect co-deviation of nodes within measurement windows, and the between-person network to reflect stable individual differences (see SI Section, “Time trend inspection and detrending and Table S5”). Other model assumptions are described in further detail elsewhere (53,60).

To reduce false-positive edges and enhance interpretability, standard pruning procedures were applied using a significance threshold of = 0.05. We assessed edge weights’ stability using bootstrapping procedures (N = 1000, SI Section “Bootstrapping procedures”). All visualized networks presented here represent pruned regularized networks to facilitate sparsity and interpretability of the most important edges and were created using the *qgraph* package (version 1.9.8; (61)). Sex-differences in network structure were tested using the Individual Network Invariance Test (INIT; (62) (SI Section “Network Invariance Testing Procedure, Table S6). Finally, the analyses were repeated, using a depression sum-score instead of specific symptoms, following the procedure described in previous network studies (42).

## RESULTS

### Descriptive Statistics

Pairwise correlations between all variables for males and females across time can be found in Table S7. Table 1 summarizes model fit indices for the saturated and the pruned models. Model fit improved slightly after pruning and the pruned model demonstrated acceptable fit in accordance with conventional cut-off criteria (CFI/TLI ≥ .89, RMSEA ≤ .05; Du et al., 2025). Extensive model fit statistics are included in Table S8. Bootstrapped estimates can be found in Figure S5-S7.

**Table 1.** Fit Indices for Full and Pruned Panel GVAR Models.

| Fit Index | Model (Full) | Model (Pruned) |
| --- | --- | --- |
| CFI | 0.89 | 0.89 |
| TLI | 0.86 | 0.88 |
| RMSEA | 0.026 | 0.024 |

*Note.* CFI = Comparative Fit Index; TLI = Tucker-Lewis Index; RMSEA = Root Mean Square Error of Approximation.

### Panel GVAR

Across all networks, the model revealed robust within-domain associations among depressive symptoms and among brain measures, with the strongest edges seen in the symptom domain (Figure 2). Significant cross-domain brain-symptom associations were observed in all networks (SI, Table S10 for overview of edge weights). Network invariance testing indicated that the structure of the within-person networks differed between males and females, whereas the between-person network structure did not significantly differ between sexes.

#### Temporal effects

The temporal network captures lagged within-person associations, reflecting whether deviations from an individual’s typical level of a symptom or brain measure predicted subsequent deviations in other variables. We observed reciprocal within-domain associations between depressed mood and worthlessness, and between lethargy and anhedonia, as well as autoregressive effects (self-loops) for all nodes, indicating temporal stability within both symptom and brain domains (e.g., higher depressed mood at one wave predicted higher depressed mood at the subsequent wave). Three significant cross-domain temporal associations emerged: higher depressed mood predicted a reduction in global surface area (r = −0.03, p = .002; bootstrap stability = 33%) and an increase in mOFC thickness (r = 0.02, p = .006; stability = 30%), and higher worthlessness predicted an increase in global surface area (r = 0.03, p = .008; stability = 27%). The modest bootstrap stability of these edges warrants cautious interpretation.

#### Contemporaneous effects

The contemporaneous network reflects within-person concurrent associations between symptoms and brain structure within the same measurement window (approximately 2 years in this study). Strong positive within-domain associations were observed, particularly among depressive symptoms (e.g., depressed mood – worthlessness, r = 0.29). One significant cross-domain contemporaneous association emerged, with higher depressed mood co-occurring with lower global surface area (r = −0.03, p < .001; bootstrap stability = 43%).

#### Between-person effects

The between-person network reflects individual differences in average levels of brain features and symptom scores. This network revealed positive within-domain associations, indicating that individuals with higher average levels on one symptom or brain measure tended to have higher average levels on others within the same domain. One significant cross-domain association emerged at the between-person level, with higher average anhedonia associated with lower average global surface area across individuals (r = −0.06, p < .001, bootstrap stability = 99.5 %).

#### Sex-specific effects

When comparing the overall network structure across all three networks simultaneously, INIT indicated that the Different model provided a superior fit relative to the Equal model (ΔAIC = +109.84, Δχ² = 571.85, df = 231, p < .001), indicating significant sex differences in global network structure. These differences were concentrated in the within-person networks, where the Different model again showed superior fit (ΔAIC = +150.93, Δχ² = 502.93, df = 176, p < .001). In contrast, no sex differences were indicated for the between-person network, where all three fit indices favoured the Equal model (ΔAIC = −44.81, Δχ² = 65.19, df = 55, p = .163), suggesting that stable trait-level brain-symptom associations are invariant across sex (Figure S7).

Visualization of pruned networks per group, showed that for the temporal effects, two negative associations from depressed mood to cortical thickness in the cingulate (*r* = -0.03) and from cortical thickness in the mOFC to lethargy (*r* = -0.03) in the male network and from global surface area to anhedonia (*r* = 0.06), worthlessness (*r* = 0.04) and depressed mood (*r* = 0.05) in the female network.

In the contemporaneous networks, negative cross-domain associations emerged only in the female network between worthlessness and fusiform gyrus thickness (*r* = -0.03) (Figure 3).

#### Depression Sum Score

Following earlier work (43), and as a sensitivity analysis, we replaced the five individual symptom nodes with the CBCL DSM-5 Depressive Problems scale (DSM_Dep_Sum, 13 items) and a self-computed five-item sum score (Dep_Sum), modelled alongside the same brain features. Both analyses yielded virtually identical results, revealing a larger number of cross-domain associations across all three networks compared to the main analysis, including temporal, contemporaneous, and between-person edges between the depression sum score and multiple cortical thickness regions, hippocampal volume, and global surface area (Figure 4 & Figure S8). Full results are reported in SI Section “Depression Sum Score.

## DISCUSSION

The present study examined within-person and between-person associations between depressive symptoms and brain structure across adolescence in a large community sample. Within persons, increases in depressed mood predicted subsequent reductions in surface area and increases in mOFC thickness, while increases in worthlessness predicted subsequent increases in surface area, indicating opposite-direction temporal associations between distinct depressive symptoms and surface area. Depressed mood was also concurrently associated with lower surface area. At the between-person level, individuals with higher average anhedonia showed stable, trait-like associations with lower surface area.

The temporal findings suggest that increases in depressed mood may be followed by reduced cortical expansion relative to an individual’s expected developmental trajectory, with the inverse pattern observed for worthlessness. Higher depressed mood also predicted increases in mOFC thickness. A parallel pattern was identified contemporaneously, though given the biennial assessment intervals and prior removal of developmental trends, these associations reflect within-person co-fluctuations within the same measurement window rather than momentary processes (51).

Although small in magnitude, these within-person findings are consistent with prior work associating surface area to depression in adolescence (16,18,22), extending these cross-sectional associations to a longitudinal within-person context. The positive temporal association from depressed mood to mOFC thickness is less straightforward to interpret, but may reflect compensatory or reactive processes in prefrontal emotion regulation regions during periods of individually elevated depressive symptoms (64). Notably, associations were driven specifically by depressed mood and worthlessness rather than anhedonia, lethargy, or sleep problems in the within-person networks. This may corroborate depressed mood’s core role in depressive symptomatology across development (65,66), and is consistent with network studies identifying depressed mood as a central symptom with high connectivity to other nodes (67).

Compared with cross-sectional symptom-level network studies in adults and older adolescents, our findings revealed fewer and weaker brain-symptom associations in a longitudinal context, which is expected given that longitudinal models estimate unique associations after removing developmental trends and accounting for stable differences. (41) reported a negative association between cingulate thickness and sadness, while (43) observed associations between worthlessness and cingulate and insula thickness in older adolescents. The absence of cingulate and fusiform associations in the present sample, which spans a younger developmental window, may reflect that distinct cortical thickness-symptom coupling in these regions emerges later in adolescence, while global surface area may be a more sensitive structural indicator of depressive symptom fluctuations in earlier adolescence.

Global network structures differed significantly by sex, with differences emerging primarily in the organization of within-person associations. In females, cross-domain associations were observed in the temporal network, indicating positive lagged coupling from global surface area to depressed mood, anhedonia and worthlessness. One negative cross-domain association emerged in the contemporaneous network from between fusiform and worthlessness. In males, cross-domain associations were confined to the temporal network, with negative associations from mOFC to lethargy and from depressed mood to cingulate. Notably, the direction and sign of some sex-specific edges diverged from those observed in the full-group model; for instance, global surface area predicted subsequent increases in depressed mood in females, whereas the pooled model showed the reverse. Such divergence may reflect true effect modification by sex, conceptually related to Simpson’s paradox (68), or partial correlation distortions if a node acts as a common effect (collider) of others in the network ((69). However, given the small effect sizes and roughly halved sample size within each sex-stratified network, reduced power offers an equally plausible explanation.

Differences in pubertal timing between sexes offer one plausible, though speculative, contributor to the observed sex differences in network organization. A sensitivity analysis examining network invariance by pubertal timing indicated that while statistically significant differences in within-person network structure were detected in both sexes, AIC consistently favored the equal-constraints model, suggesting that pubertal timing differences are smaller in practical magnitude than the sex differences identified in the primary analysis (Figure S14). Future work incorporating pubertal development is needed to clarify this further (70).

Aggregating depressive symptoms into either the full CBCL Depressive Problems score (13 items) or a self-computed five-item sum score revealed a larger number of cross-domain associations across within-person and between-person networks, contrasting with (42) finding that granular symptom-brain associations are obscured by sum score aggregation.

Key strengths of the present study include the large longitudinal sample and the separation of within-person dynamics from stable between-person differences. Several design features likely contribute to the subtle effect sizes observed, including detrending of developmental trajectories, estimation of partial correlations isolating unique associations, a community sample with relatively low symptom severity, and multi-year assessment intervals (71). The findings are consistent with growing evidence that reproducible brain–behavior associations in large samples tend to be modest (72). Given the small effect sizes and pronounced floor effects in the symptom distributions, individual edge weights should be interpreted with caution.

Several limitations should be considered. First, the panel GVAR framework estimates sample-average networks and cannot capture individual-specific or subgroup-level dynamics, potentially obscuring meaningful heterogeneity (53,60). Second, brain regions of interest were selected a-priori, enhancing comparability with previous studies (41,43), but potentially overlooking distributed effects or effects that may emerge using other imaging modalities, such as diffusion-weighted imaging (73,74). Third, depressive symptoms were assessed using parent-reported CBCL items, which may not fully capture internalizing symptoms as adolescents age, given well-established declines in parent-child agreement across adolescence (75,76). A sensitivity analysis in a clinically enriched subsample yielded broadly similar network structures (Figure S12). Fourth, the community-based sample likely reflects subclinical symptom levels with limited variance that may constrain the detectability of associations and yield conservative edge weight estimates. Future studies should examine whether these associations replicate in symptom-enriched or clinical samples, and whether these associations persist after accounting for time-varying confounders such as medication use and adverse life events, as the current within-person design, while removing time-invariant confounding, cannot rule out these dynamic influences. Fifth, the biennial spacing of assessment waves constrains interpretation, as depressive episodes are typically shorter in duration (77). Future studies employing denser sampling designs may be better positioned to resolve shorter-term neurobiological-affective coupling and its developmental timing (78,79). Finally, partial correlation networks are sensitive to the set of included nodes, and results may vary with different symptom items or brain features. Replication across samples with varied node selection will be important for establishing the robustness of the present findings.

## Conclusion

In sum, this study provides evidence that associations between depressive symptoms and brain structure in adolescence are subtle, symptom-specific, and present at both the within- and between-person level. These findings highlight the value of longitudinal, symptom-level approaches for capturing both trait-like brain correlates of depressive symptoms and the dynamic, within-person coupling between mood and brain structure across development.

## Data Availability

Data from the Adolescent Brain Cognitive Development (ABCD) Study are publicly available to qualified researchers through the NIMH Data Archive (NDA), featuring longitudinal neuroimaging, behavioral, and genetic data. Access requires an approved Data Use Certification (DUC).

https://doi.org/10.82525/ab4q-qr87

## ACKNOWLEDGEMENTS

This work was supported by the Research Council of Norway (#288083, #323951) and the South-Eastern Norway Regional Health Authority (#2021070, #2023012, #500189). Data was handled inside Service for Sensitive Data (TSD), a platform owned by the University of Oslo, operated, and developed by the TSD service group at the University of Oslo IT-Department (USIT). Computations were performed using resources provided by UNINETT Sigma2 – the National Infrastructure for High Performance Computing and Data Storage in Norway (NS9666S). Data used in the preparation of this article were obtained from the <u>Adolescent Brain Cognitive Development™ (ABCD) Study</u>, held in the <u>NIH Brain Development Cohorts Data Sharing Platform</u>. This is a multisite, longitudinal study designed to recruit more than 10,000 children aged 9–10 and follow them over 10 years into early adulthood.

The ABCD Study® is supported by the National Institutes of Health and additional federal partners under award numbers:

U01DA041048, U01DA050989, U01DA051016, U01DA041022, U01DA051018, U01DA05 1037, U01DA050987, U01DA041174, U01DA041106, U01DA041117, U01DA041028, U01 DA041134, U01DA050988, U01DA051039, U01DA041156, U01DA041025, U01DA041120, U01DA051038, U01DA041148, U01DA041093, U01DA041089, U24DA041123, U24DA041147. A full list of supporters is available at <u>Federal Partners – ABCD Study</u>. ABCD Consortium investigators designed and implemented the study and/or provided data but did not necessarily participate in the analysis or writing of this report. This manuscript reflects the views of the authors and may not reflect the opinions or views of the NIH or ABCD Consortium investigators. A version of this article is published as a preprint on medRxiv and is currently undergoing screening.

## FIGURE LEGENDS

**Figure 1. Age distributions split by sex and timepoint.**

*Note*. Timepoint 1 represents baseline data, and timepoints 2–4 represent 2-year, 4-year, and 6-year follow-up assessments. Females are shown in purple and males in green. Dotted lines connect sex-specific mean ages across timepoints. Error bars represent ± standard error of the mean.

**Figure 2. Temporal (A), contemporaneous (B), and between-person (C) pruned networks.**

Note. Blue nodes represent depressive symptoms: anhedonia (Anhed.), worthlessness (Worthless.), lethargy, depressed mood (Dep. Mood), and sleep problems (Sleep). Pink nodes represent structural brain metrics: fusiform gyrus, insula, cingulate cortex, medial orbitofrontal cortex (mOFC), hippocampus (Hippoc.), and global surface area (Global SA). Blue edges indicate positive partial correlations and pink edges indicate negative partial correlations. Only edges surviving the pruning step (α = .05) are shown. Edge labels reflect partial correlation coefficients.

**Figure 3. Sex-specific temporal (A, C) and contemporaneous (B, D) networks**

*Note*. Male networks (A, B): light green nodes represent depressive symptoms and dark green nodes represent structural brain metrics. Female networks (C, D): light purple nodes represent depressive symptoms and dark purple nodes represent structural brain metrics. Blue edges indicate positive partial correlations between nodes and red edges indicate negative partial correlations. Edge transparency is uniform across all edges regardless of magnitude. Between-person networks are presented in Supplementary Figure S7.

**Figure 4. Temporal (A), contemporaneous (B), and between-person (C) pruned networks from the depression sum score sensitivity analysis.**

Note. Networks estimated using an aggregated CBCL DSM-5 Depressive Problems scale (DSM_Dep_Sum, 13 items) as a single node, modelled alongside the same six brain features as the main analysis: fusiform gyrus, insula, cingulate cortex, medial orbitofrontal cortex (mOFC), hippocampus (Hippoc.), and global surface area (Global SA). A parallel analysis using a self-computed five-item sum score (Dep_Sum) yielded virtually identical results (Supplementary Figure S8). The blue node represents the depression sum score and pink nodes represent structural brain metrics. Blue edges indicate positive partial correlations and pink edges indicate negative partial correlations. Only edges surviving the pruning step (α = .05) are shown. Edge labels reflect partial correlation coefficients.

